# Hypertension Management Dynamics in Pediatric CKD: Insights from the 4C Study

**DOI:** 10.1101/2024.11.18.24317519

**Authors:** Anke Doyon, Aysun Karabay Bayazit, Ali Duzova, Daniela Thurn, Nur Canpolat, Ipek Kaplan Bulut, Karolis Azukaitis, Lukasz Obrycki, Bruno Ranchin, Rukshana Shroff, Cengiz Candan, Hakan Erdogan, Dusan Paripovic, Osman Donmez, Francesca Lugani, Klaus Arbeiter, Ebru Yilmaz, Ariane Zaloszyc, Elke Wühl, Anette Melk, Uwe Querfeld, Franz Schaefer, 4C Study Consortium

## Abstract

**Rationale:** To investigate blood pressure (BP) trajectories, and the impact of pharmacological intervention in children with chronic kidney disease (CKD).

**Methods:** Analysis of antihypertensive treatment (AHT) and BP slopes in 320 patients of the 4C Study cohort with CKD prior to renal replacement therapy, based on a minimum of three individual observations and two years of follow-up.

**Results:** At enrollment, 70 patients (22%) had uncontrolled or untreated hypertension, 130 patients (41%) had controlled hypertension, and 120 patients (37%) had normotension without antihypertensive treatment. AHT medication was prescribed in 53% of patients at baseline and initiated or added in 91 patients (AHT-I, 28%) during follow-up.

Overall BP standard deviation score (SDS) remained stable over time in the cohort (ß= -0.037±0.034, p=0.34 and -0.029± 0.348, p=0.093 per year for systolic and diastolic BP SDS). In the AHT-I group, systolic and diastolic BP SDS was higher at baseline and decreased significantly during follow-up (−0.22±0.07, p<.003 and -0.12±0.05 SDS per year, p=0.01). Only 8/70 (11%) patients of the previously untreated/uncontrolled group remained untreated at the last observation, while 31 (44%) were controlled during follow-up. Of the 120 normotensive patients at baseline, 60% remained normotensive while 40% progressed to uncontrolled/untreated (n=23, 19%) or controlled (n=24, 20%) hypertension.

**Conclusions:** The study provides comprehensive real-world evidence on long-term management of blood pressure in children with CKD from the 4C Study. Although blood pressure control improved significantly with the intensification of antihypertensive therapy, a notable proportion of previously normotensive patients developed de novo hypertension over the observation period.

## Introduction

Hypertension represents a cardinal risk factor for the progression of chronic kidney disease (CKD) and the development of cardiovascular disease (CVD). It is omnipresent among patients with CKD with a significant variation in prevalence across different degrees of renal dysfunction^1–3^. While optimizing BP is therefore one of the hallmarks of controlling secondary complications of CKD^4^, previous studies have highlighted the challenges in achieving this goal in pediatric and adult patients^1,3,5,6^.

Few studies and trials have examined the blood pressure dynamics and the impact of antihypertensive treatment on actual BP control in children with CKD over extended periods of time, especially in a real-world setting^7^. The aim of this study was to describe blood pressure classification and antihypertensive therapy longitudinally, using data from the Cardiovascular Comorbidity in Children with Chronic Kidney Disease - 4C - Study cohort^8^. Specifically, the analysis examined individual antihypertensive medication prescriptions and their impact on blood pressure trends over time.

## Methods

### Patients and Study Design

All patients were enrolled in the 4C (Cardiovascular Comorbidity in Children with Chronic Kidney Disease) Study, which was conducted in 55 pediatric nephrology units across 12 European countries. The study design and objectives have been described previously in detail^8^. The study prospectively observed 704 patients with chronic kidney disease aged 6-17 years with an estimated glomerular filtration rate below 60 ml/min/1.73m^2^ and no kidney replacement therapy (KRT) at the time of inclusion. The study population was enrolled between 2009 and 2012. During this period, annual regional investigator-led anthropometric measurements, evaluations of kidney function, blood pressure (BP) assessments, and cardiovascular monitoring were performed. Furthermore, anthropometric and blood pressure measurements were conducted at six-month intervals by local investigators. A comprehensive medication history was documented for each patient.

The 4C study was approved by the Ethics Committee of Heidelberg University (S-032/2009) and the institutional review boards at each participating institution. Written informed consent was obtained from all parents and participants as appropriate. The study was registered at ClinicalTrials.gov on August 7, 2009, with the identifier NCT01046448). The data and methods used in the analysis will be made accessible to any researcher for the purpose of reproducing the results or replicating the procedures, in accordance with the AHA Journals’ Implementation of the Transparency and Openness Promotion (TOP Guidelines).

The baseline visits, in addition to two available consecutive annual follow-up visits conducted by a regional study coordinator, were included for descriptive analysis if they had valid laboratory analysis and BP measurements. Consequently, patients who had both a baseline visit and a complete one- and two-year follow-up, as well as patients who had a complete baseline visit, one-year follow-up, and three-year follow-up, but missed the second-year follow-up, were also eligible for inclusion in the descriptive analysis. For the purposes of longitudinal analysis, all available BP measurements from interim visits, conducted at six-months intervals between the initial and final selected visit were additionally included, even in the absence of laboratory values at the time. The visits were classified according to the sequence of the selected visits conducted by the regional study investigators and designated as baseline, first follow-up, and last follow-up, respectively. Body mass index standard deviation score (BMI SDS) and height standard deviation score (height SDS) were calculated using the reference data provided by the World Health Organization (WHO).^9^

### Office BP Measurements and classification of hypertension

Office BP measurements were performed using oscillometric devices validated for pediatric use with appropriate cuff sizes. The median of three consecutive measurements was used for analysis. Systolic and diastolic BP were standardized to SD scores (SDS) by accounting for age, height and sex^10^.

Hypertension was defined as systolic or diastolic (BP) above the 95th percentile for age, height, and sex for patients below 16 years of age and BP greater or equal to 140/90 mmHg for patients above 16 years^4^. The target ranges for BP, defined in accordance with the 2016 European Society of Hypertension guidelines, were a BP SDS below the 75th percentile for patients without proteinuria and the 50th percentile with proteinuria in patients below the age of 16^4^. In patients above the age of 16 years, the cut-off values were defined as 130/80 mmHg in the absence of proteinuria and 125/75 mmHg in the presence of proteinuria.

### Medication

Medication records were analyzed by extracting all medications used for blood pressure control at the selected yearly visits. BP medications were grouped according to mechanism of action into inhibitors of the renin-angiotensin-system (RAS Inhibitors: ACE or AT1 blockers), calcium channel blockers, beta blockers, alpha blockers, loop diuretics and other diuretics. For a detailed list of prescribed medications, see supplementary material.

Intensification of antihypertensive (AHT) medication was defined by the addition of another AHT medication with an alternative mechanism of action, or by the initiation of any AHT medication when no AHT treatment was performed before. Patients with intensification of AHT medication will henceforth be referred to as the antihypertensive intensification group (AHT-I) as opposed to patients without start or adding of AHT medication (AHT-N).

A sensitivity analysis was conducted to determine the impact of dose adjustments by mg per body weight in kg between annual visits. The analysis identified groups based on the escalation of AHT therapy, either through the addition of AHT medication or the increase of the dose of the prevalent AHT medication. The analysis demonstrated that dosage increases based on body weight were typically minimal or did not result in a net increase until the conclusion of the observation period. Consequently, we postulated that patients who initiated or added antihypertensive drugs were deemed to have undergone a notable intensification of antihypertensive treatment, whereas those who experienced isolated dose increases did not. Further details regarding this analysis can be found in the supplementary material, including Table S3, Figure S3, and Figure S4.

### Data analysis

Descriptive statistics were calculated for all yearly visits, stratified by visit number, hypertension status, and depending on the intensification of AHT medication during follow-up. The data are presented as mean (SD), median (IQR), or percentage, as appropriate. Between-group comparisons were analyzed using Kruskal-Wallis rank sum test and Pearson’s Chi-squared test, as appropriate. Descriptive comparisons between first and last observations were carried out by paired t-test for continuous variables and McNemar test for nominal variables. For comparison across multiple time points, the Friedman test as a non-parametric test and repeated reasures ANOVA for continuous variables was used. A linear mixed model for repeated measurements was fitted for both absolute and age-, height- and sex-adjusted systolic and diastolic blood pressure measurements, adjusting for baseline BP values and including random intercepts per patient and, when the model allowed it, also random slopes per patient. Contrast statements were formulated to assess the significance of differences between the intercepts and slopes depending on AHT intensification. All figures with slopes display a time frame until 3 years after baseline, since only few patients had their last included yearly visit at a later time point.

P values <0.05 were considered statistically significant. Data analysis and the generation of figures was conducted using SAS version 9.3 (SAS Institute) and R Studio ^11^.

## Results

Basic characteristics of the selected 320 patients are shown in Table 1. eGFR decreased significantly during follow-up from 30±10 to 24±11 ml/min/1.73 m^2^.

**Table 1.** Basic characteristics at baseline and annual follow-up visits. Data are given as mean (SD) or n (%).

| Follow Up | Baseline<br>visit<br>N = 320<sup>1</sup> | 1<sup>st</sup> Follow-up<br>N = 320<sup>1</sup> | Last Follow-up<br>N = 320<sup>1</sup> | P value** |
| --- | --- | --- | --- | --- |
| Age | 10.9 (2.9) | 12.1 (2.9) | 13.2 (2.9) | <0.001 |
| Male gender |  | 211 (66%) |  |  |
| BMI SDS | 0.02 (1.30) | 0.11 (1.14) | 0.06 (1.28) | 0.1647 |
| eGFR (ml/min/1.73 m<sup>2</sup>) | 29.9 (10.1) | 27.5 (10.8) | 24.2 (11.2) | <0.001 |
| **Blood pressure** |  |  |  |  |
| Systolic BP (mmHg) | 109.4 (13.5) | 110.6 (14.4) | 112.4 (14.7) | 0.001 |
| Diastolic BP (mmHg) | 67.1 (11.6) | 66.5 (10.4) | 67.8 (11.5) | 0.1517 |
| Systolic BP SDS | 0.76 (1.3) | 0.66 (1.25) | 0.63 (1.27) | 0.205 |
| Diastolic BP SDS | 0.62 (1.01) | 0.46 (0.90) | 0.49 (1.00) | 0.027 |
| **Hypertension status\*** |  |  |  | 0.043 |
| Normotension | 120 (37.5%) | 113 (35.3%) | 102 (31.9%) |  |
| Untreated hypertension | 31 (9.7%) | 21 (6.6%) | 28 (8.8%) |  |
| Controlled hypertension | 130 (40.6%) | 137 (42.8%) | 152 (47.5%) |  |
| Uncontrolled hypertension | 39 (12.2%) | 49 (15.3%) | 38 (11.9%) |  |
| BP within recommended range* | 84 (26%) | 114 (36%) | 106 (33%) | 0.01 |
| BP below 95th percentile | 250 (78%) | 250 (78%) | 254 (79%) | 0.872 |
| **Antihypertensive medication** | 169 (53%) | 186 (58%) | 190 (59%) | 0.012 |
| **Number of AHT medications** |  |  |  | 0.006 |
| 0 | 151 (47%) | 134 (42%) | 130 (41%) |  |
| 1 | 118 (37%) | 123 (38%) | 131 (41%) |  |
| 2 | 35 (11%) | 45 (14%) | 41 (13%) |  |
| ≥ 3 | 16 (5%) | 18 (6%) | 18 (6%) |  |
| **Medication Type** |  |  |  |  |
| RAS Inhibitors | 131 (41%) | 137 (43%) | 130 (41%) | 0.584 |
| Calcium channel antagonists | 47 (15%) | 70 (22%) | 69 (22%) | <0.001 |
| Beta Blockers | 26 (8%) | 23 (7%) | 34 (11%) | 0.027 |
| Alpha Blockers | 8 (3%) | 15 (5%) | 15 (5%) | 0.03 |
| Loop diuretics | 18 (6%) | 17 (5%) | 15 (5%) | 0.417 |
| Other diuretics | 11 (3%) | 10 (3%) | 11 (3%) | 0.717 |

\* according to European Society of Hypertension Guidelines 2014<sup>4</sup>, \*\*Repeated measures Anova for continuous and Friedman test for categorical variables

### Blood pressure at baseline and during follow-up

Systolic and diastolic BP SDS values remained stable (ß=-0.037 ±0.034, p=0.34 and ß=-0.029 ±0.020, p=0.14 per year). The overall prevalence of normotension without AHT medication was 37% at time of enrollment, and 32% at last observation (p=0.09). The fraction of patients with controlled hypertension increased from 41% at baseline to 48% at last follow-up (p=0.028). The prevalence of uncontrolled or untreated hypertension was 22% at baseline and remained unchanged throughout the study period. **(Table 1, Figure 2)**.

**Figure 1:**
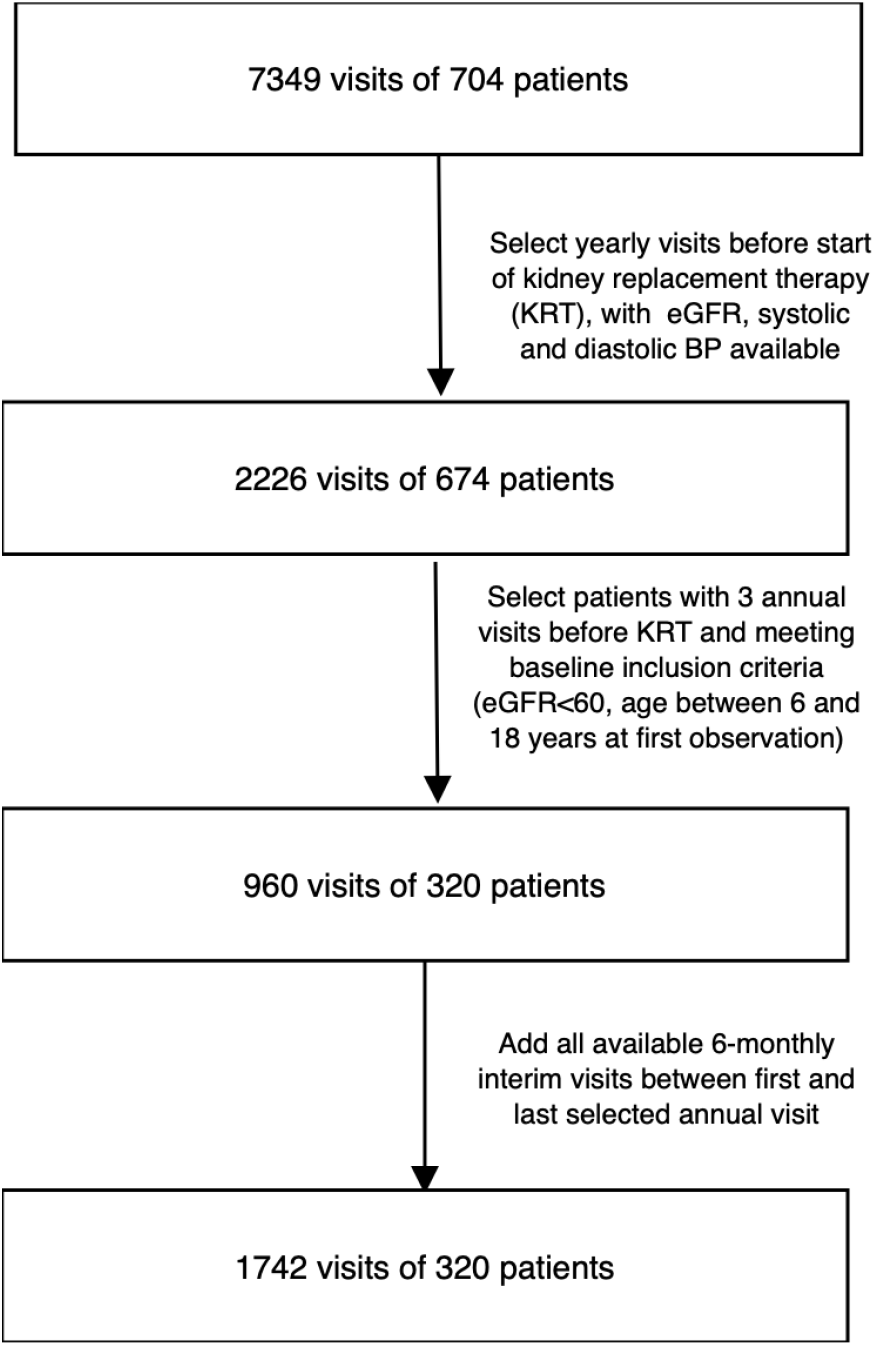
Patient and visit selection

**Figure 2:**
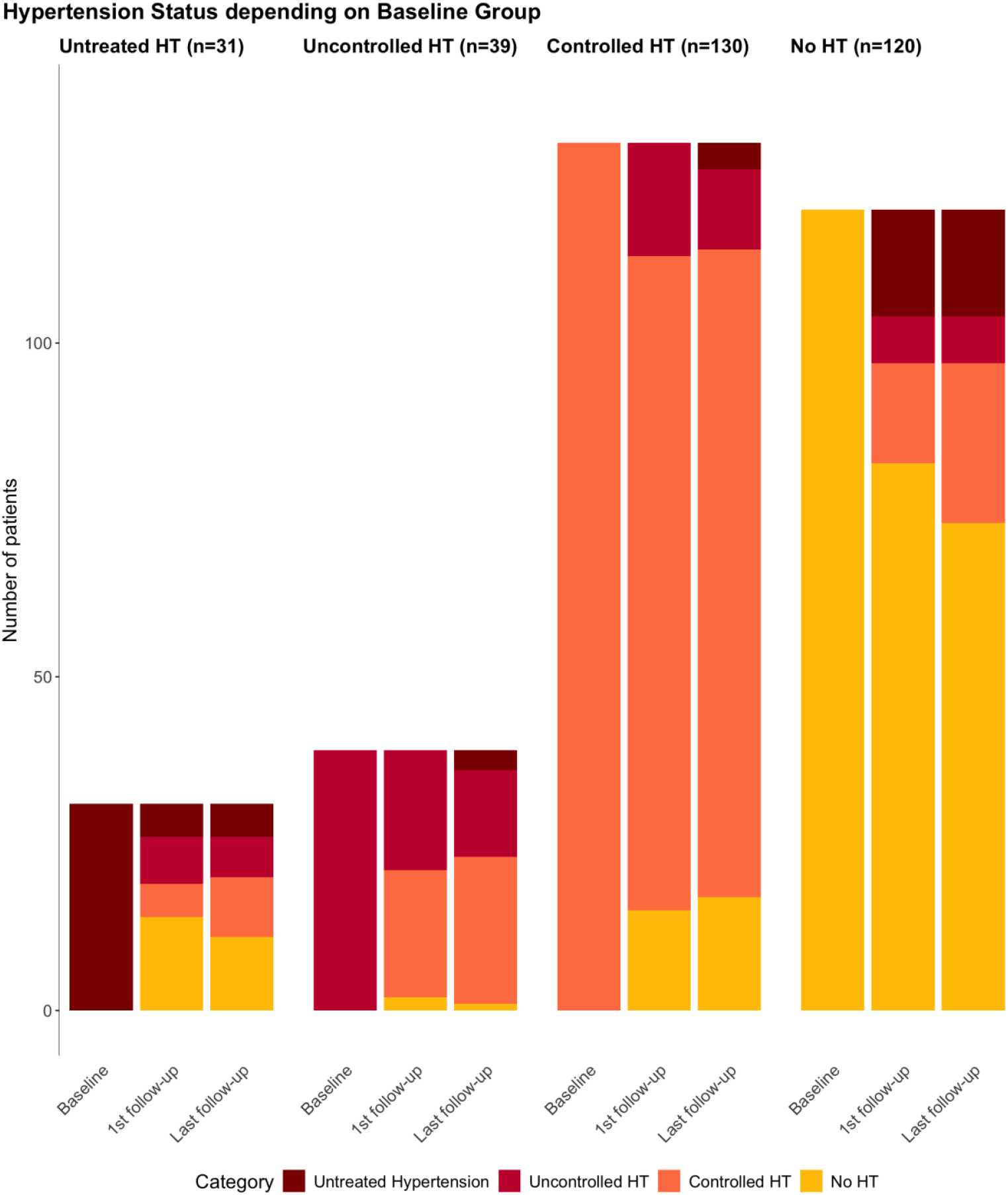
Hypertension status during annual follow-up visits according to baseline blood pressure status.

Among the patients with any hypertension at baseline, antihypertensive medication was initiated or augmented in 38 patients (32%) with initially controlled hypertension and in 31 patients (44%) with initially untreated/uncontrolled HT at baseline (**Table S2b**).

Among 70 patients with untreated or uncontrolled hypertension at baseline, 44.3% achieved a controlled hypertensive state, while 38.6% remained either untreated (n=8, 11.4%) or uncontrolled (n=19, 27.1%). Additionally, 17% exhibited normotensive blood pressure without the use of antihypertensive medication at the final follow-up. Conversely, a significant number of patients (n=39, 15.6%) from the normotensive/untreated and controlled hypertension groups progressed to an uncontrolled or untreated hypertensive state during the observation period (**Figure 2, Table S2a**). In **longitudinal modeling** of systolic and diastolic BP SDS, BP SDS decreased significantly during follow-up in patients with previously uncontrolled or untreated patients but remained stable in patients who were normotensive or had controlled hypertension at baseline (**Figure S2**).

### Impact of intensified AHT medication

In the entire cohort, 91 patients (28%) were started on a first or additional antihypertensive medication during the observation period (‘AHT-I’-group, **Figure 3, Table 2)**.

**Table 2.**
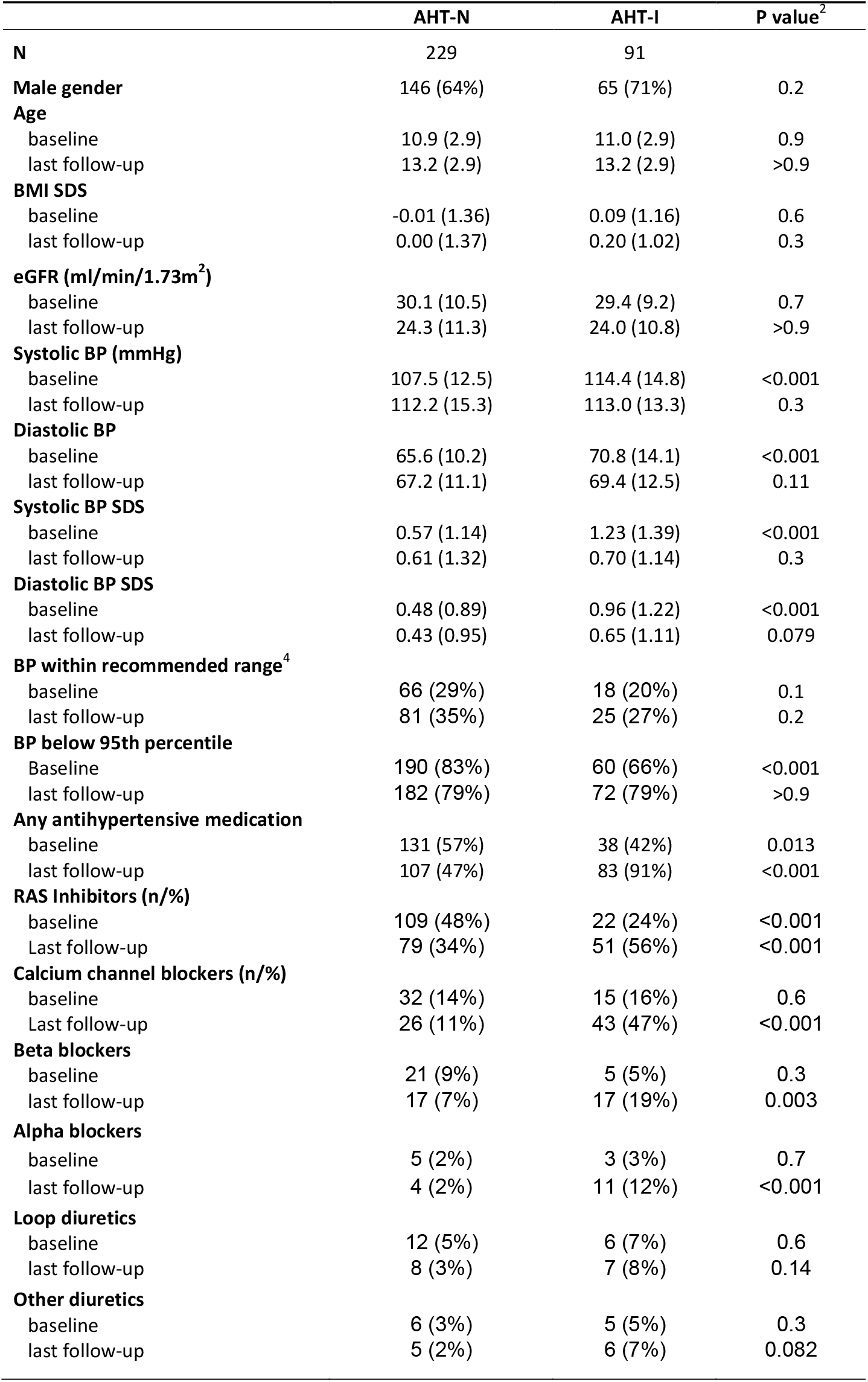
Cohort characteristics at first and last visit according to change in antihypertensive medication. **AHT-I**: Patients with added AHT medication or commencing AHT, **AHT-N**: Patients without new or any AHT medication during follow-up. Data are given as mean (SD) and n (%).

|  | AHT-N | AHT-I | P value<sup>2</sup> |
| --- | --- | --- | --- |
| **N** | 229 | 91 |  |
| **Male gender** | 146 (64%) | 65 (71%) | 0.2 |
| **Age** |  |  |  |
| baseline | 10.9 (2.9) | 11.0 (2.9) | 0.9 |
| last follow-up | 13.2 (2.9) | 13.2 (2.9) | >0.9 |
| **BMI SDS** |  |  |  |
| baseline | -0.01 (1.36) | 0.09 (1.16) | 0.6 |
| last follow-up | 0.00 (1.37) | 0.20 (1.02) | 0.3 |
| **eGFR (ml/min/1.73m<sup>2</sup>)** |  |  |  |
| baseline | 30.1 (10.5) | 29.4 (9.2) | 0.7 |
| last follow-up | 24.3 (11.3) | 24.0 (10.8) | >0.9 |
| **Systolic BP (mmHg)** |  |  |  |
| baseline | 107.5 (12.5) | 114.4 (14.8) | <0.001 |
| last follow-up | 112.2 (15.3) | 113.0 (13.3) | 0.3 |
| **Diastolic BP** |  |  |  |
| baseline | 65.6 (10.2) | 70.8 (14.1) | <0.001 |
| last follow-up | 67.2 (11.1) | 69.4 (12.5) | 0.11 |
| **Systolic BP SDS** |  |  |  |
| baseline | 0.57 (1.14) | 1.23 (1.39) | <0.001 |
| last follow-up | 0.61 (1.32) | 0.70 (1.14) | 0.3 |
| **Diastolic BP SDS** |  |  |  |
| baseline | 0.48 (0.89) | 0.96 (1.22) | <0.001 |
| last follow-up | 0.43 (0.95) | 0.65 (1.11) | 0.079 |
| **BP within recommended range<sup>4</sup>** |  |  |  |
| baseline | 66 (29%) | 18 (20%) | 0.1 |
| last follow-up | 81 (35%) | 25 (27%) | 0.2 |
| **BP below 95th percentile** |  |  |  |
| Baseline | 190 (83%) | 60 (66%) | <0.001 |
| last follow-up | 182 (79%) | 72 (79%) | >0.9 |
| **Any antihypertensive medication** |  |  |  |
| baseline | 131 (57%) | 38 (42%) | 0.013 |
| last follow-up | 107 (47%) | 83 (91%) | <0.001 |
| **RAS Inhibitors (n/%)** |  |  |  |
| baseline | 109 (48%) | 22 (24%) | <0.001 |
| Last follow-up | 79 (34%) | 51 (56%) | <0.001 |
| **Calcium channel blockers (n/%)** |  |  |  |
| baseline | 32 (14%) | 15 (16%) | 0.6 |
| Last follow-up | 26 (11%) | 43 (47%) | <0.001 |
| **Beta blockers** |  |  |  |
| baseline | 21 (9%) | 5 (5%) | 0.3 |
| last follow-up | 17 (7%) | 17 (19%) | 0.003 |
| **Alpha blockers** |  |  |  |
| baseline | 5 (2%) | 3 (3%) | 0.7 |
| last follow-up | 4 (2%) | 11 (12%) | <0.001 |
| **Loop diuretics** |  |  |  |
| baseline | 12 (5%) | 6 (7%) | 0.6 |
| last follow-up | 8 (3%) | 7 (8%) | 0.14 |
| **Other diuretics** |  |  |  |
| baseline | 6 (3%) | 5 (5%) | 0.3 |
| last follow-up | 5 (2%) | 6 (7%) | 0.082 |

**Figure 3.**
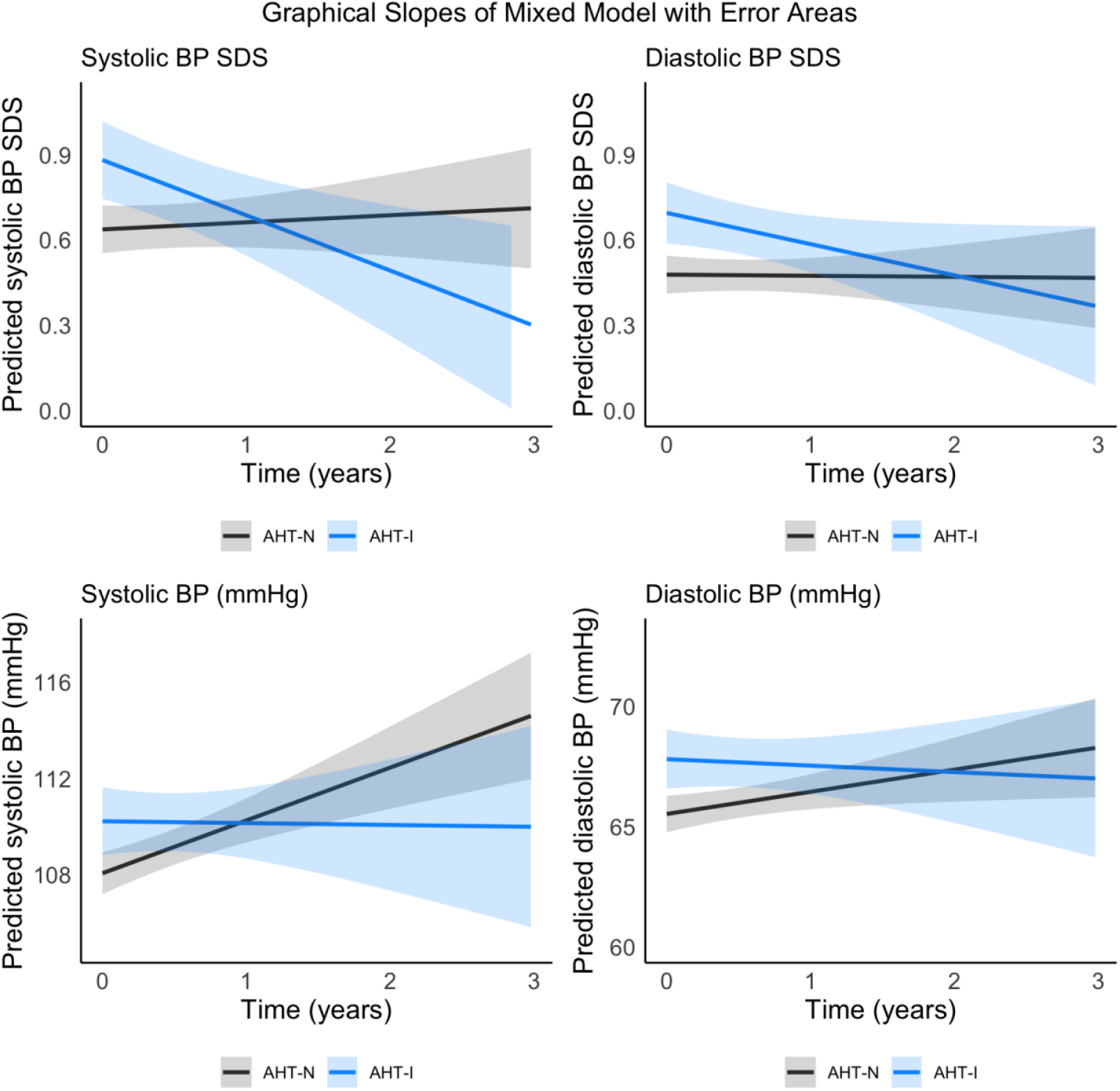
Systolic and diastolic blood pressure slopes in patients with initiated or intensified AHT therapy (AHT-I, blue) compared to patients with unchanged treatment status (AHT-N, grey). Upper panels: BP SDS, lower panels: absolute BP values. Lines and shaded areas represent mean slopes and error areas.

Patients in whom antihypertensive medication was started or intensified (AHT-I group) had higher systolic and diastolic BP at baseline (systolic BP SDS 1.23±1.39 vs 0.57±1.14, p<0.001) but their BP did not differ from the other patients (AHT-N group) at the last follow-up (systolic BP z-score 0.7±1.14 vs 0.61±1.32, p=0.3). The distribution of sex, age, BMI SDS and renal function did not differ between both groups. For additional characteristics of the AHT-I and AHT-N groups, see **Table 2**.

The BP slope analysis revealed no significant changes in overall BP z-scores over time (‘Time since baseline’). However, patients in the AHT-I group exhibited a significant decline in both systolic and diastolic BP SDS, with an average annual reduction of -0.22 ± 0.07 for systolic BP SDS (p = 0.003) and - 0.12 ± 0.05 for diastolic BP SDS (p = 0.01). Model contrasts further demonstrated that patients in the AHT-I group had significantly elevated BP at baseline compared to those in the AHT-N group, as well as a more pronounced decline in BP over time (**Table 3, Figure 3**).

**Table 3:** Longitudinal mixed model describing impact of intensified antihypertensive treatment (AHT-I) on standardized systolic and diastolic blood pressure.

|  | Systolic BP Z-score |  |  | Diastolic BP Z-score |  |  |
| --- | --- | --- | --- | --- | --- | --- |
|  | Estimate ± SE | 95% CI | p | Estimate ± SE | 95% CI | p |
| Intercept | 0.14 ± 0.05 | 0.05 - 0.23 | 0.002 | 0.19 ± 0.05 | 0.10 - 0.28 | <0.001 |
| BP SDS at baseline | 0.66 ± 0.03 | 0.61 - 0.72 | <0.001 | 0.45 ± 0.03 | 0.39 - 0.51 | <0.001 |
| Time since baseline | 0.02 ± 0.04 | -0.05 - 0.10 | 0.529 | 0.00 ± 0.02 | -0.04 - 0.05 | 0.968 |
| AHT-I group | 0.24 ± 0.08 | 0.08 - 0.41 | 0.003 | 0.30 ± 0.08 | 0.13 - 0.46 | <0.001 |
| Time since baseline for AHT-I group | -0.22 ± 0.07 | -0.36 - -0.07 | 0.003 | -0.12 ± 0.05 | -0.20 - -0.03 | 0.010 |
| **Contrasts for AHT-I yes vs. no** |  |  |  |  |  |  |
|  |  | **Chisq** | **p** |  | **Chisq** | **p** |
| **Intercept** |  | 8.774 | 0.003 |  | 12.47 | <0.001 |
| **Slope** |  | 8.635 | 0.003 |  | 6.656 | 0.0099 |

Regarding absolute blood pressure (BP) values, there was a notable increase in both systolic and diastolic BP in the AHT-N group over the course of the study, whereas systolic BP exhibited a significant decline in the AHT-I group (Supplementary material).

In the AHT-I group, 31 (34%) patients had untreated or uncontrolled hypertension at baseline and 24 (26%) at last follow-up. 68 patients (75%) in the AHT-I group were normotensive at baseline and 77 patients (85%) at last follow-up (p=0.11). The number of patients who met the ESH guideline-recommended BP targets increased from 84 (26%) to 106 (33%) (p<0.031).

### Prescription of antihypertensive medication

The prevalence of antihypertensive medication (AHT) prescription was 53% (n=169) at baseline and 59% (n=190) at last observation. The majority of patients received monotherapy throughout the observation period (37% at baseline and 41% at last follow-up), while only 16% at baseline and 19% at last follow-up were treated with combined AHT therapy. Triple therapy was prescribed only in a minority of patients (5-6%). The most frequently prescribed medications were RAS inhibitors (41-43% of patients), with no significant change throughout the study period. The use of calcium channel blockers and beta blockers increased significantly between over time (15% to 22% of patients, p=0.001 and 8-11%, p=0.027), whereas diuretics and alpha-blockers were constantly used in 8-9% and 3-5% of patients respectively throughout follow-up (Table 1, Figure 4).

**Figure 4.**
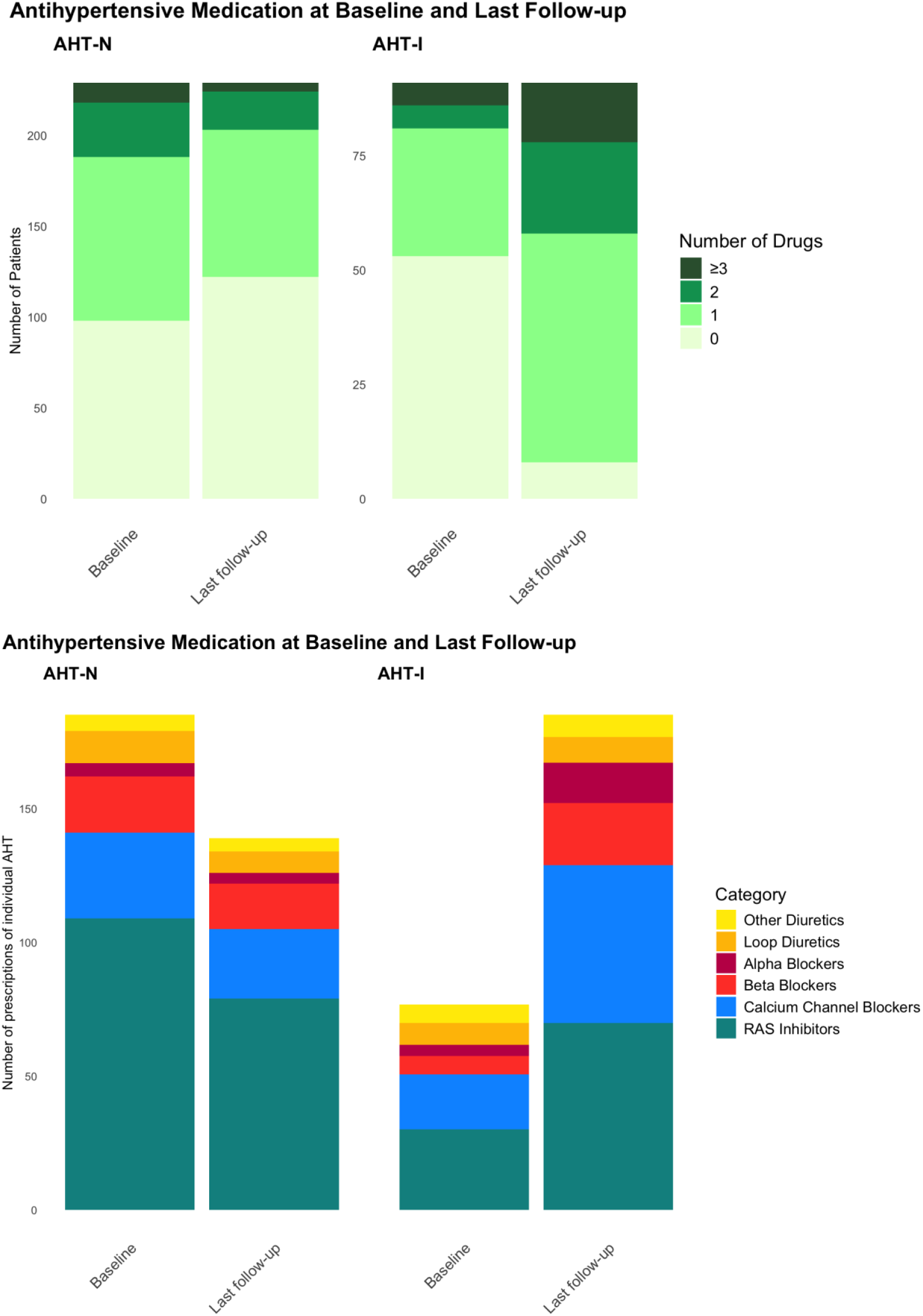
Antihypertensive medication at first and last visit in patients with and without intensified therapy. Upper panel: Number of antihypertensive drugs per patient. Lower panel: Cumulative number of individual prescriptions by class.

In the AHT-I group, the overall number of prescriptions for any AHT medication increased by 141% with an increase of 132% for the prescription of RAS inhibitors and 186% for calcium antagonists. (Table 2, Figure 4). Conversely, the use of AHT prescriptions decreased by 25% in the AHT-N group, with RAS inhibitors decreasing by 28% and calcium channel blockers by 19%.

Combination therapy was more common in patients of the AHT-I group at last follow-up (38%) compared to baseline (11%) and compared to the AHT-N group at any time (18% at baseline and 11% at last follow-up) (Figure 4).

## Discussion

This analysis of the 4C Study describes blood pressure trajectories and longitudinal patterns of antihypertensive treatment in a large group of pediatric patients with CKD in a real-life setting, yielding important new insights.

In this longitudinal study in children with CKD stage 3-5 not on kidney replacement therapy, a constant fraction of around 21% of patients were hypertensive at any point in time and mean age-standardized blood pressure did not change over time. This observation confirms previous findings in the North American CKiD cohort, where casual BP in the hypertensive range were constantly observed in 14% of children with slightly less advanced CKD^12^. In the KNOW-PedCKD study of Korean children with CKD stage 1-5, overall hypertension prevalence was 31% at baseline^13^.

However, our analysis stratified by hypertension status at baseline revealed that the unchanged blood pressure status over time at the cohort level is the net result of dynamic, reciprocal changes in blood pressure evolution and management that occurred in different subgroups of the cohort. Among the patients with untreated hypertension at baseline, 48% received antihypertensive treatment at last follow-up. Control of hypertension was achieved in more than half of the patients with uncontrolled hypertension at baseline. On the other hand, more than a third of the initially normotensive patients developed hypertension during follow-up, 50% of whom were controlled by AHT medication whereas 13% remained untreated.

Antihypertensive therapy was prevalent in 53% of patients at time of enrolment, slightly less frequently than in the CKiD and HOT-KID study cohorts where the baseline prevalence of AHT medication was 64% and 65% respectively^16,5^. In contrast to the extensive published evidence for the high prevalence of hypertension in children with CKD, there is little information regarding the efficacy of antihypertensive therapy under real-world conditions. Even among interventional trials, few studies followed patients for sufficiently long periods of time to assess blood pressure slopes. A recent Cochrane analysis identified only 13 trials with more than 50 pediatric patients monitored for ≥ 4 weeks ^14^.

We observed a significant and sustained improvement of BP control in patients in whom antihypertensive therapy was started or intensified during the observation period. Systolic and diastolic BP decreased by approximately 0.2 SD per year in these patients and the proportion of patients meeting the ambitious recommended ESH guideline targets increased from 26 to 33%. While these changes were significant, our findings show that real-world blood pressure management is less effective than what can be achieved in clinical trial settings. For comparison, in the ESCAPE trial overall blood pressure level dropped by approximately 1 SD within 6 months of the antihypertensive intervention and was maintained around the 50^th^ percentile for up to six years of follow-up^15^.

The majority of patients in our cohort were prescribed RAS antagonists at some point, in line with current recommendations. The overall prescription rate of RAS inhibitors remained stable during the observation time. But while in the AHT-I group there was a steep increase of prescription of RAS inhibitors, the reduction in antihypertensive medication in the AHT-N group was mainly accounted for by discontinuation of RAS inhibitors. While the reasons for discontinuation of AHT were not recorded, progression of CKD and associated adverse effects may have contributed to the discontinuation specifically of RAS inhibitors.

Only few patients (5-7% in the AHT-I group) were treated with diuretics. This could be due to the paucity of clinical trials with diuretics in children and to concerns regarding efficacy and safety of diuretic treatment. In contrast, loop diuretics and thiazides are frequently used for treatment in adult CKD patients and recent evidence supports their beneficial effect ^16,17^. Treatment with chlortalidone may effectively contribute to blood pressure control even in advanced CKD stages^18^. Studies in children with CKD should clarify whether diuretic treatment in children with CKD is safe and effective, and could provide additional therapeutic options for better control of hypertension.

Our findings demonstrate that a considerable fraction of children with CKD remain undertreated despite the availability of pediatric evidence-based clinical practice guidelines. The majority of patients in this study were prescribed monotherapy throughout the observation period. While first-line monotherapy is recommended in the current pediatric ESH guideline ^4^, the 2024 ESH clinical practice guideline for the management of hypertension^19^ in adults recommends the initiation of antihypertensive treatment with a combination of two drugs, i.e. a RAS antagonist plus a calcium channel blocker or a diuretic. Undertreatment of hypertension as well as other comorbidities in children with CKD has been shown to be highly prevalent in several studies ^12,20^, seemingly without improvement during follow-up^2^. We here show that this seems to be in part attributable to delayed treatment of *de novo* hypertension that occurs in patients with progressive CKD. Furthermore, even when treatment is initiated or intensified not all patients achieve the desired blood pressure goals. Earlier antihypertensive therapy – maybe even in prehypertensive states - may help to curb the prevalence of undertreated hypertension. Moreover, aggressive intensification of AHT could increase the successful achievement of BP goals.

Our analysis is based on an observational cohort. Since office BP measurements are frequently used in daily routine for adjusting blood pressure medication, office BP measurements were exclusively used for this analysis. How this translates in more elaborate blood pressure patterns from ambulatory blood pressure monitoring and the prevalence of masked hypertension will be subject to subsequent analysis. Although there was a significant decline in eGFR in the study cohort, the patients selected for this analysis were relatively stable, as a minimum of three visits with at least annual intervals before starting renal replacement therapy were required to meet the eligibility criteria. Therefore, the results may not be generalizable to patients who are closer to starting kidney replacement therapy. In our analysis, dosage increases were only minor in most cases, therefore it is not possible to determine from this study whether the administration of effective dose adjustments in addition to or before drug escalation would have resulted in improved outcomes. However, this is a question that should be considered in both our daily practice and in future clinical trials.

**In conclusion**, this is the first study describing real world patterns of antihypertensive treatment and its effect on BP over time in a large cohort of children with CKD. Antihypertensive treatment was initiated or intensified in a substantial number of patients, suggesting that treatment guidelines are implemented by clinicians. The observed treatment intensification was effective to improve blood pressure control. Dynamic changes in response to treatment modifications and progression of CKD with new-onset hypertension resulted in unchanged overall blood pressure SDS and prevalence of hypertension over time. There is an evident need for strategies to curb the overall prevalence of hypertension in children with CKD.

## Data Availability

The data and methods used in the analysis will be made accessible to any researcher for the purpose of reproducing the results or replicating the procedures, in accordance with the AHA Journals? Implementation of the Transparency and Openness Promotion (TOP Guidelines).

## Acknowledgements

Support for the 4C Study was received from the ERA-EDTA Research Program, the KfH Foundation for Preventive Medicine and the German Federal Ministry of Education and Research (01EO0802). The study was also supported by the European Reference Network for Rare Kidney Diseases (ERKNet), which is funded by the European Union within the framework of the EU4Health Program (101085068).

